# CNM-Au8 Treatment Is Associated With Reduced Chronic Lesion Tissue Expansion in Relapsing-Remitting Multiple Sclerosis

**DOI:** 10.64898/2026.09.23.26363841

**Authors:** Samuel Klistorner, Michael Hotchkin, Ben Greenberg, Austin Rynders, Robert Glanzman, Heidi Beadnall, Steve Vucic, Richard MacDonell, Jeannette Lechner-Scott, William Carroll, Stefan Blum, Anneke Van der Walt, Bruce Taylor, Deborah Field, Michael Barnett, Alexandr Klistorner

## Abstract

**Background:** CNM-Au8, a catalytic nanocrystalline gold suspension, is proposed to support neuronal bioenergetics, but its effect on chronic lesion evolution in multiple sclerosis (MS) has not been reported.

**Objectives:** To evaluate the effect of CNM-Au8 on chronic lesion tissue expansion (CLTE).

**Results:** In this exploratory post hoc analysis of the VISIONARY randomised placebo-controlled trial, CLTE was quantified from serial FLAIR MRI. Fifty-five participants were included (CNM-Au8, n=40; placebo, n=15). Compared with placebo, CNM-Au8 was associated with lower lesion expansion rate (p=0.001; Cliff’s δ=™0.58) and volume (p<0.001; Cliff’s δ=™0.65).

**Conclusions:** CNM-Au8 treatment was associated with substantially reduced chronic lesion tissue expansion.

## Introduction

Progressive disability accumulation in multiple sclerosis (MS) is increasingly recognised as being driven not only by acute inflammatory lesion formation, but also by chronic pathological processes within established white matter lesions. A subset of chronic lesions continues to expand slowly over time, reflecting ongoing tissue destruction at lesion boundaries associated with persistent microglial activation, demyelination and axonal injury. Chronic lesion tissue expansion (CLTE), quantified on longitudinal magnetic resonance imaging (MRI), has been associated with accelerated brain atrophy and disability progression.[1] CLTE has therefore emerged as a promising imaging biomarker of chronic destructive lesion activity in MS.[2]

CNM-Au8 is a suspension of catalytic nanocrystalline gold particles proposed to enhance cellular bioenergetics and support neuroaxonal function.[3][4][5] Although not developed specifically to target chronic inflammatory lesion activity, improved metabolic resilience and repair capacity may plausibly influence processes underlying chronic lesion expansion.

We performed an exploratory blinded post hoc imaging analysis of the VISIONARY trial to determine whether CNM-Au8 treatment was associated with reduced CLTE in relapsing MS.

## Methods

This was an exploratory post hoc imaging analysis of the VISIONARY randomised placebo-controlled trial evaluating CNM-Au8 in relapsing MS. Study design and inclusion criteria can be found elsewhere.[6]

Serial fluid-attenuated inversion recovery (FLAIR) images were co-registered longitudinally. Lesion masks were generated on FLAIR images using semi-automated segmentation with blinded lesion analysis. Participants with a baseline chronic lesion volume <100mm3 were excluded from analysis.

CLTE was quantified using a validated in-house lesion progression algorithm implemented in Python, as previously described. [1] Briefly, lesions were matched across timepoints based on spatial overlap, while new free-standing and confluent lesions were identified and excluded from CLTE analysis. Follow-up lesion masks were spatially adjusted to account for atrophy-related displacement before lesion volume change was calculated. CLTE was expressed as annualised percentage change (CLTE rate, %/year) and as annualised absolute volumetric change (mm^3^/year). Placebo-treated participants were followed until the end of the double-blind phase, whereas CNM-Au8-treated participants were followed until their last available MRI obtained during the open-label extension.

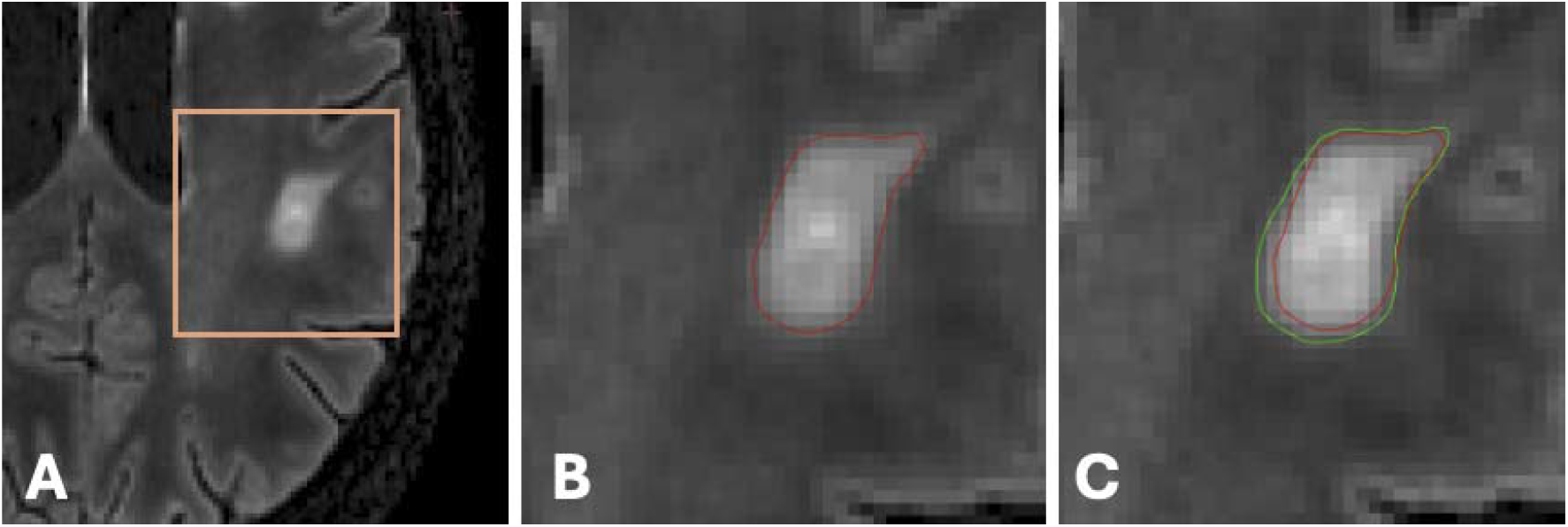

## Statistical analysis

Within-group longitudinal change was assessed using a one-sample t-test with Cohen’s d calculated as the standardised mean change. Between-group comparisons were performed using the Mann-Whitney U test, with Cliff’s delta (δ) calculated as a standardised effect size. Effect sizes were interpreted as negligible (<0.15), small (0.15–0.33), medium (0.33–0.47), or large (>0.47). To address a trend imbalance in disease duration between groups (p=0.055), a sensitivity analysis was performed using ordinary least-squares regression adjusted for disease duration, using a signed-log–transformed outcome and heteroscedasticity-consistent standard errors (HC3). Statistical significance was defined as p<0.05. All analyses were performed in Python.

## Results

### Study population

Imaging data were available for 74 participants. Two participants without follow-up MRI, eight with <6 months of follow-up, and nine with baseline chronic lesion volume <100 mm^3^ were excluded, leaving 55 participants for analysis (placebo, *n* = 15; CNM-Au8, *n* = 40).

Baseline demographic and clinical characteristics were generally comparable between groups (Table 1). Disease duration showed a trend toward shorter duration in the CNM-Au8 group (p=0.055).

**Table 1.** Baseline demographic and clinical characteristics. IQR = interquartile range. P-values reflect Placebo vs Treatment (both doses combined): Mann-Whitney U for continuous variables; Chi-square for categorical variables. ^ EDSS data unavailable for 2 treatment subjects (n=38). DMT category: H = high-potency; I = intermediate-potency; L = low-potency.

| Variable | Placebo (n=15) | Treatment (n=40) | P-value |
| --- | --- | --- | --- |
| Age (yrs), median [IQR] | 37 [33, 39.5] | 39.5 [31.8, 47.0] | 0.603 |
| Sex (F:M) | 12:3 | 27:13 | 0.565 |
| BMI, median [IQR] | 26.98 [24.6, 32.5] | 25.77 [23.0, 29.5] | 0.380 |
| Disease duration (yrs), median [IQR] | 7.0 [5.0, 8.5] | 4.0 [1.0, 7.25] | 0.055 |
| EDSS $\leq 1.5$ / $>1.5$ | 8 / 7 | 20 / 18 ^ | 1.000 |
| DMT category, n | H:8, I:5, L:1, Nil:1 | H:24, I:11, L:1, Nil:4 | 0.834 |
| Baseline lesion volume (mm<sup>3</sup>), median [IQR] | 3,442 [416, 5,434] | 1,187 [491, 3,046] | 0.261 |

### Chronic lesion expansion

#### Within-group longitudinal change

In the placebo group, mean annual chronic lesion volume increased significantly during follow-up (3,892 ± 3,723 mm^3^ vs 4,273 ± 3,989 mm^3^ at baseline and follow-up respectively; p=0.0001), representing a mean absolute change of +381 ± 287 mm^3^ (Cohen’s d = 1.33). There was also a significant increase in mean chronic lesion volume in the treatment group (mean 2,737 ± 4,089 mm^3^ vs 2,852 ± 4,054 mm^3^ at baseline and follow-up respectively, p<0.0001). A mean absolute change was +114 ± 224 mm^3^, Cohen’s d = 0.51).

#### Between-group comparison

Compared with placebo, CNM-Au8 treatment was associated with significantly lower chronic lesion expansion across both outcome measures (Fig. 2). Median expansion rate was 8.4%/year in the placebo group versus 2.7%/year in the treatment group (p=0.001; Cliff’s δ = −0.58, 95% CI −0.83 to −0.30). Median expansion volume was 311 mm^3^/year in the placebo group versus 14 mm^3^/year in the treatment group (p<0.001; Cliff’s δ = −0.65, 95% CI −0.88 to −0.36). A sensitivity analysis adjusting for disease duration, which showed a trend toward imbalance between groups (p=0.055), did not materially alter these findings.

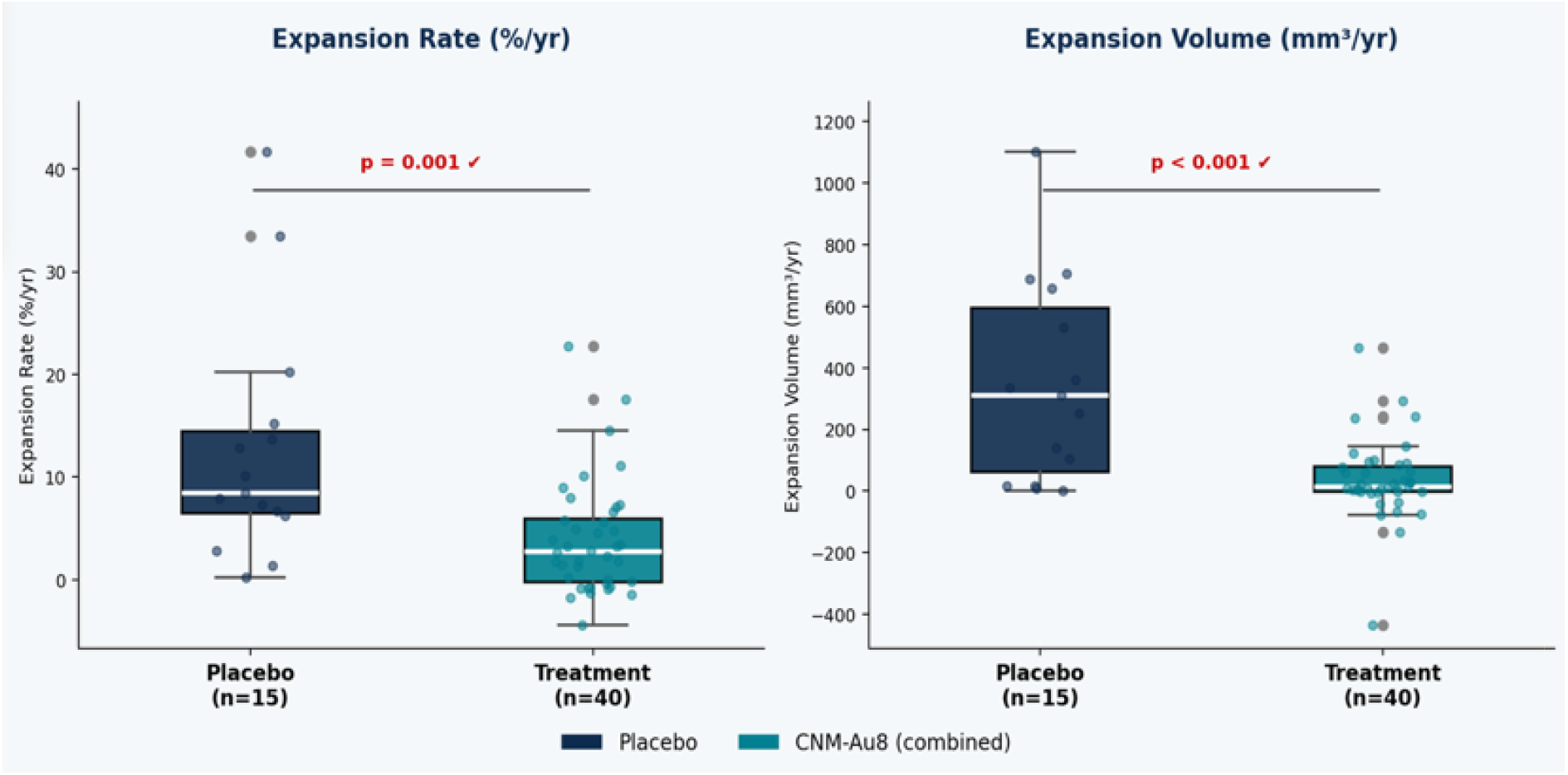

## Discussion

In this blinded exploratory post hoc imaging analysis of the VISIONARY trial, CNM-Au8 treatment was associated with a marked reduction in chronic lesion tissue expansion in relapsing MS. The effect was consistent across non-parametric analyses and covariate-adjusted regression models.

Chronic lesion expansion is thought to reflect persistent tissue injury at lesion boundaries, where microglial activation, oxidative stress, mitochondrial dysfunction, oligodendrocyte injury and impaired remyelination may create a state of sustained metabolic vulnerability.[2][7][8][9] These processes may perpetuate gradual tissue destruction and lesion enlargement over time. This provides a plausible biological context for the observed effect of CNM-Au8. Rather than acting as a conventional anti-inflammatory therapy, CNM-Au8 is proposed to support cellular energy metabolism by facilitating NADH oxidation to NAD+ and increasing ATP availability.[10] Human target-engagement studies using 7T phosphorus magnetic resonance spectroscopy have demonstrated increased brain NAD+/NADH ratios following treatment, supporting a CNS bioenergetic effect in vivo.[4] Experimental studies also suggest effects on oligodendrocyte differentiation and remyelination.[10] Enhancement of cellular bioenergetic capacity within metabolically stressed lesion rims could therefore improve tissue resilience and repair, shifting lesion evolution away from progressive tissue destruction towards stabilisation. The reduction in CLTE observed here is consistent with such a mechanism, although it does not establish it.

Several limitations should be acknowledged. This was a post hoc exploratory analysis with a relatively small sample size, particularly in the placebo arm. Follow-up was longer and more variable in the CNM-Au8 group because treatment continued into the open-label extension, whereas placebo follow-up ended with the double-blind phase. Although outcomes were annualised, an effect of follow-up duration cannot be excluded. CLTE was not a predefined trial endpoint, dose groups were combined because all participants crossed to 30 mg in the open-label extension, and no direct pathological or molecular measures of chronic inflammation were available. Nevertheless, the consistency and magnitude of the observed effect support further investigation of CNM-Au8 for modifying chronic lesion evolution in MS.

## Data Availability

All data produced in the present study are available upon reasonable request to the authors

